# X-ray measurement of periarticular soft tissue predict readmission complications after Total Knee Arthroplasty

**DOI:** 10.1101/2024.03.24.24304790

**Authors:** Hanwen Hu, Ye Tao, Zheng Qingyuan, Guoqiang Zhang, Ming Ni

**Author notes:** Guoqiang Zhang is the corresponding author. Hanwen Hu and Ye Tao are co-first authors.

## Abstract

**Purpose:** Obesity is widely recognized as one of the risk factors for osteoarthritis. This study aims to explore the association between BMI and periarticular soft tissue on readmission complications in patients with previous joint replacement through the study of clinical and imaging data.

**Methods:** This retrospective study summarized 625 patients who underwent total knee arthroplasty. Imaging measurement data included several measurable soft tissue values and ratios on the anteroposterior. The association between BMI and imaging soft tissue measurement data with complications leading to readmission during follow-up was explored.

**Results:** Analyzing 761 preoperative imaging measurements and postoperative follow-up data, a significant correlation (P<0.05) emerged between postoperative readmission complications and measured soft tissue thickness around the joint. A 1:4 paired test affirmed the independent predictive power (P<0.1) of select soft tissue data for readmission complications.

**Conclusion:** BMI proved insufficient in evaluating obesity-related complications post total knee arthroplasty. Preoperative imaging soft tissue data exhibited superior predictive capability for anticipating readmission complications after arthroplasty.

Trial registration number: S2021-094-01

Trial registration dates: 25/03/2021 Retrospectively registered

## Background

Osteoarthritis (OA) is the most common degenerative joint disease in middle-aged and older people [1]. Total knee arthroplasty, as the most mature surgery of the 20th century, is an effective means of treating end-stage knee osteoarthritis. However, postoperative complications have always been a concern for joint surgeons. In the past 10 years, the incidence of postoperative complications after total knee arthroplasty has not effectively decreased [3]. Taking more measures to curb the incidence of postoperative complications may increase unnecessary treatment for many low-risk patients, requiring additional costs while having little effect on them. If patients can be stratified for risk in advance and personalized treatment measures are formulated, it will be beneficial to the prognosis of some high-risk patients [4].

Previous studies have shown that age, gender, obesity and previous joint injury are well-known risk factors for knee OA [1], with obesity being closely related to the incidence of knee OA [5]. However, whether obesity is an independent risk factor for postoperative complications after total knee arthroplasty is a matter of debate among many studies. Obesity affects surgical outcomes in many ways, including systemic inflammation [6], increased weight load, increased difficulty and time of surgery [7], but many reports have expressed that BMI has not shown good efficacy in predicting postoperative complications in patients [8,9]. The possible reason is that BMI simply reflects the relationship between patient weight and height. Although modern medicine considers obesity to be a state of fat layer accumulation and low skeletal muscle mass/function [10], studies on obesity related to knee replacement often involve BMI [11], which may lead to a gap between clinical research results and physiological expectations.

Measuring the soft tissue around the knee joint to estimate the degree of obesity in patients and further predict the prognosis of knee total knee arthroplasty has begun to be studied [12]. The existing results of such experiments are also not uniform. Although most measurements of soft tissue around the knee joint show some degree of association with patient outcomes [13,14], the selection of soft tissue measurement locations used in each experiment is relatively arbitrary and not the same. Some research results believe that thicker soft tissue may be a protective factor for postoperative complications [15], and even some research results believe that BMI may have better predictive performance [16]. Although fat accumulation has a negative effect on wound healing due to its impact on systemic inflammation, some studies have shown that local fat and soft tissue thickness measurements can avoid some confounding factors of BMI [14].

This experiment statistically analyzed total knee replacement surgeries performed at our hospital from January 2019 to December 2020 and followed them up for 20 to 44 months. Clinical, BMI and imaging data were collected for patients. The patient’s BMI and preoperative anteroposterior and lateral views of the femur, patella and tibia around the soft tissue measurement data were observed. The association between patient BMI and preoperative periarticular soft tissue thickness with postoperative readmission complications was explored.

## Materials and Methods

A retrospective case-control study approved by the institutional review board was conducted among patients who had undergone total knee arthroplasty at our hospital. The clinical data and soft tissue data on preoperative anteroposterior and lateral x-rays of the knee joint collected before knee total knee arthroplasty were compared.

### Cases

We reviewed the clinical databases of multiple medical centers at our hospital and collected total knee replacement surgeries performed at all medical centers from January 2019 to December 2020. The age of patients at the time of reoperation was limited to 65-80 years to reduce the impact of age on knee prognosis. As a result, we collected a total of 1263 total knee replacements performed on 1045 patients and followed them up for 20 to 44 months, with a follow-up deadline of August 2022. Incomplete medical records, incomplete height and weight data that could not be used to calculate BMI data, and patients who could not be tracked during follow-up were excluded. The remaining 964 patients underwent 1141 total knee replacements. The imaging data of the patients were examined. Patients participating in data collection were required to have anteroposterior and lateral views of the knee joint collected within one year before surgery at the corresponding medical center. The images could be recognized and accurately measured in the Any image viewing system or Uniweb image viewing system. All anteroposterior and lateral views of the knee joint had markers to accurately adjust the zoom ratio of the image to accurately measure the thickness of bones and related soft tissues. All patients who did not meet the above imaging requirements could not be included in the study. At this point, we still had a total of 761 total knee replacements performed on 625 patients included in the study. [Figure 1]

**Figure 1.**
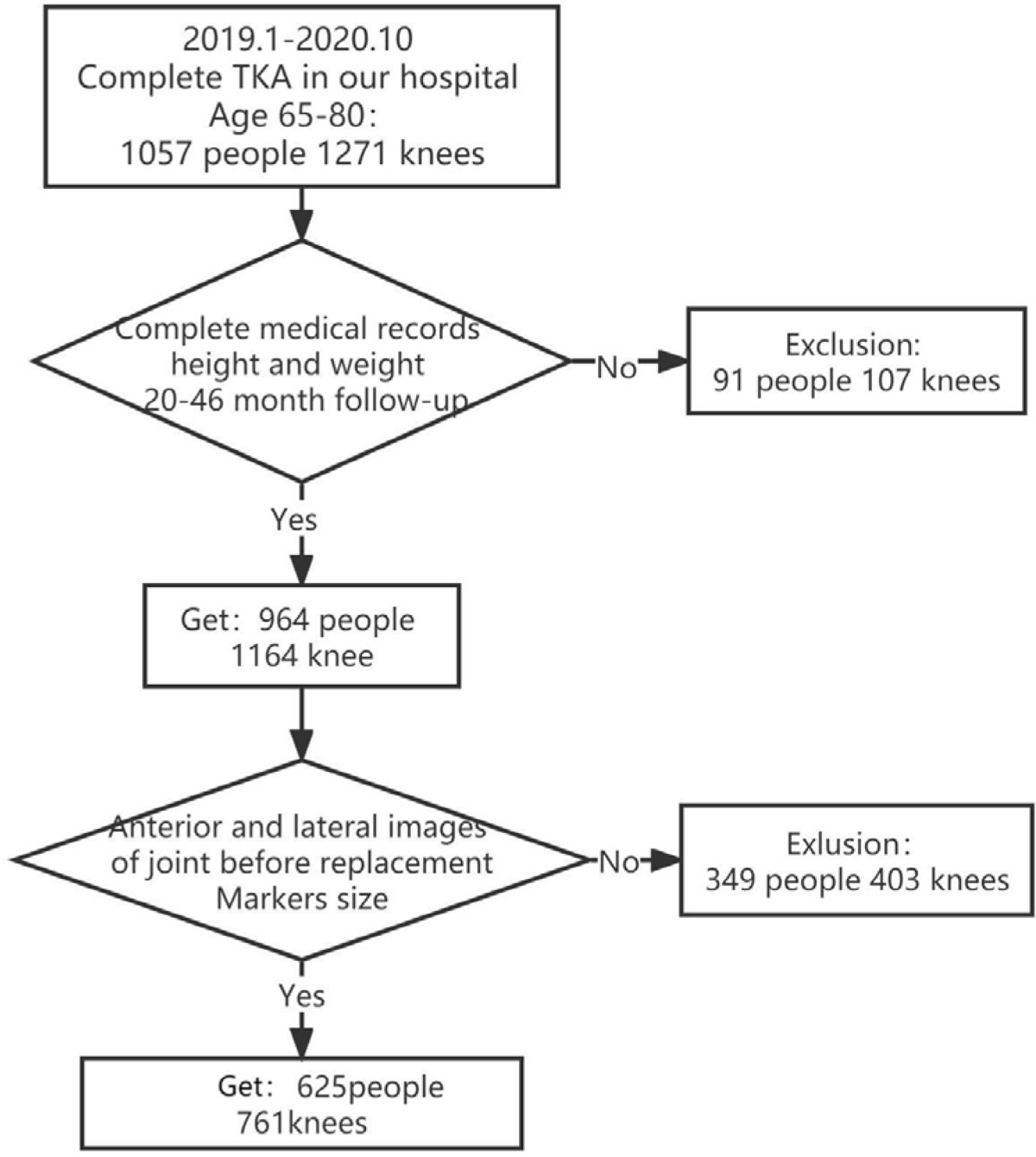
Flow diagram of the selecting process in the study

### Control

All patients included in the study underwent total knee replacement surgery performed by joint surgeons with more than 10 years of experience and who had completed more than 150 total knee replacements. All patients included in the study successfully completed the surgery and were discharged after meeting the discharge requirements. 1. Collect patient characteristics at admission, including gender, height, weight, past medical history, ASA score, side of surgery, etc. 2. Collect the patient’s age at surgery and imaging data taken within one year before surgery. The imaging data should include measurable anteroposterior and lateral views of the patient’s knee joint. The image size is calibrated using a 30mm or 50mm marker recorded on the image when the image is taken. The patient’s knee joint is flexed at 10°-40° when taking the lateral view. The Medcom AnyPacs2.0 system or EBM technologies Uniweb Server system can be used for accurate measurement.

The thickness of bones and related soft tissues is accurately measured. The measurements include: 1 Anterior femur subcutaneous tissue thickness: On the lateral view of the knee joint, locate the upper edge of the femoral condyle and measure 3cm upward along the upper edge of the femoral condyle to locate the measurement section. Measure the distance from the surface of the quadriceps to the surface of the skin on the measurement section and define it as the thickness of the anterior femur subcutaneous tissue. 2 Anterior tibia soft tissue thickness: On the lateral view of the knee joint, find the tibial tuberosity and measure the distance from the surface of the tibial tuberosity to the skin, defined as anterior tibia soft tissue thickness. 3 Antepatellar soft tissue thickness: On the lateral view of the knee joint, locate the midpoint position on the anterior surface of the patella and draw a perpendicular line through it. Measure the distance from this perpendicular line from the midpoint on the surface of patella to skin and define it as antepatellar soft tissue thickness. 4 Patella bone thickness: There are many bone spurs on patella, but bone spurs only play a stabilizing role in joint movement in their extension area. Therefore, on lateral view of knee joint, measure solid thickness at midpoint on anterior surface of patella. 5 Medial knee subcutaneous tissue thickness: On anteroposterior view of knee joint, locate midpoint on medial side of femoral condyle as plane and measure distance from upper surface of medial collateral ligament to skin on medial side of knee joint on this plane, defined as medial knee subcutaneous tissue thickness. 6 Femoral condyle width: On anteroposterior view of knee joint, remove bone spurs from femoral condyles and measure line between midpoint on inner edge of medial femoral condyle and midpoint on outer edge of lateral femoral condyle, defined as width between two femoral condyles. 7 Antepatellar soft tissue ratio: The ratio of antepatellar soft tissue thickness to patellar bone thickness on left or right lower limb image of patient. 8 Medial knee tissue to femoral condyle ratio: The ratio of medial knee subcutaneous tissue thickness to femoral condyle width on left or right lower limb image of patient. [Figure 2] The first six imaging data are measured for each patient and then calculated to obtain latter two ratios. 3. Collect postoperative readmission and reoperation data for complications with follow-up periods between 20 and 44 months with a follow-up record deadline of July 31, 2022. Patients who were diagnosed with postoperative complications or other diseases directly related to total knee arthroplasty after readmission during follow-up; patient readmission status, reoperation status and patient death status (all known follow-up patients survived). If a patient has readmission and reoperation related to joint again, record clinical diagnosis of postoperative complications in patient. If there are multiple postoperative readmissions events, they should also be recorded.

**Figure 2.**
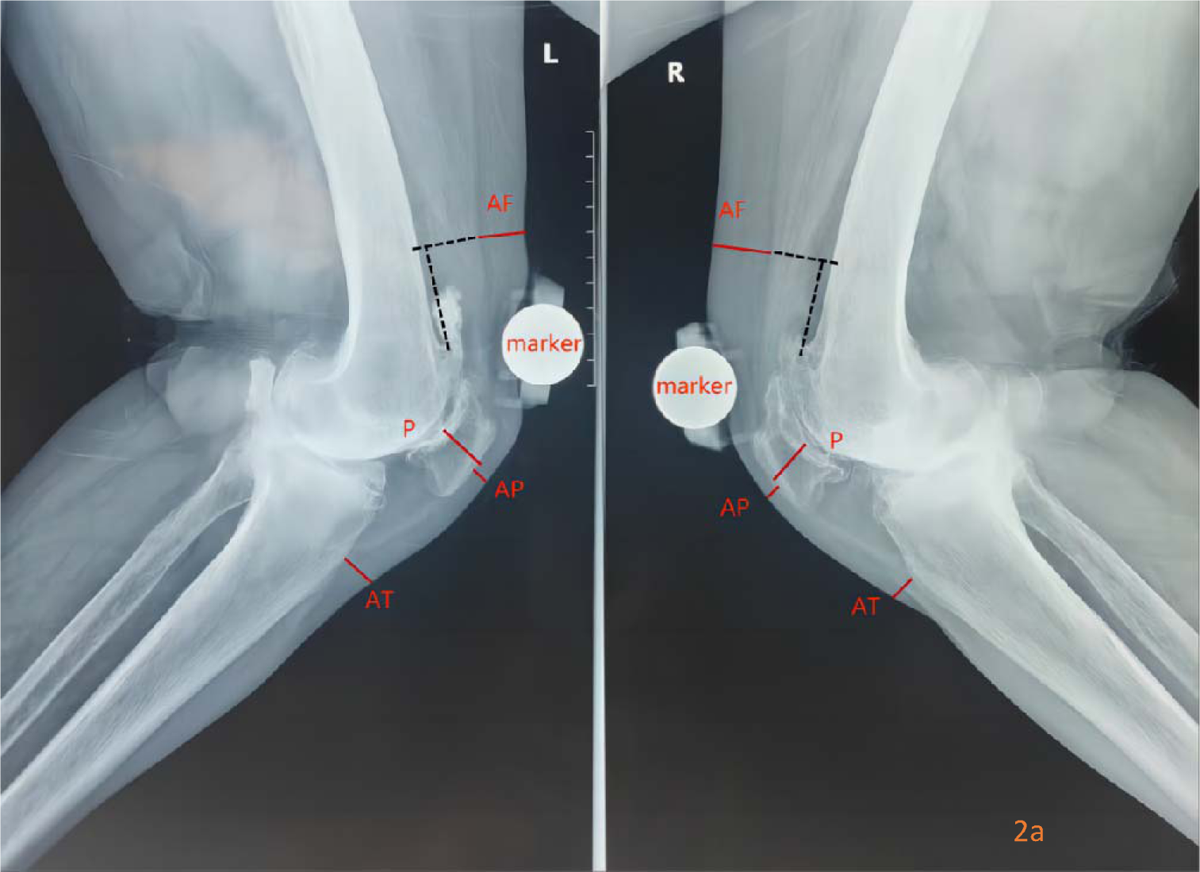
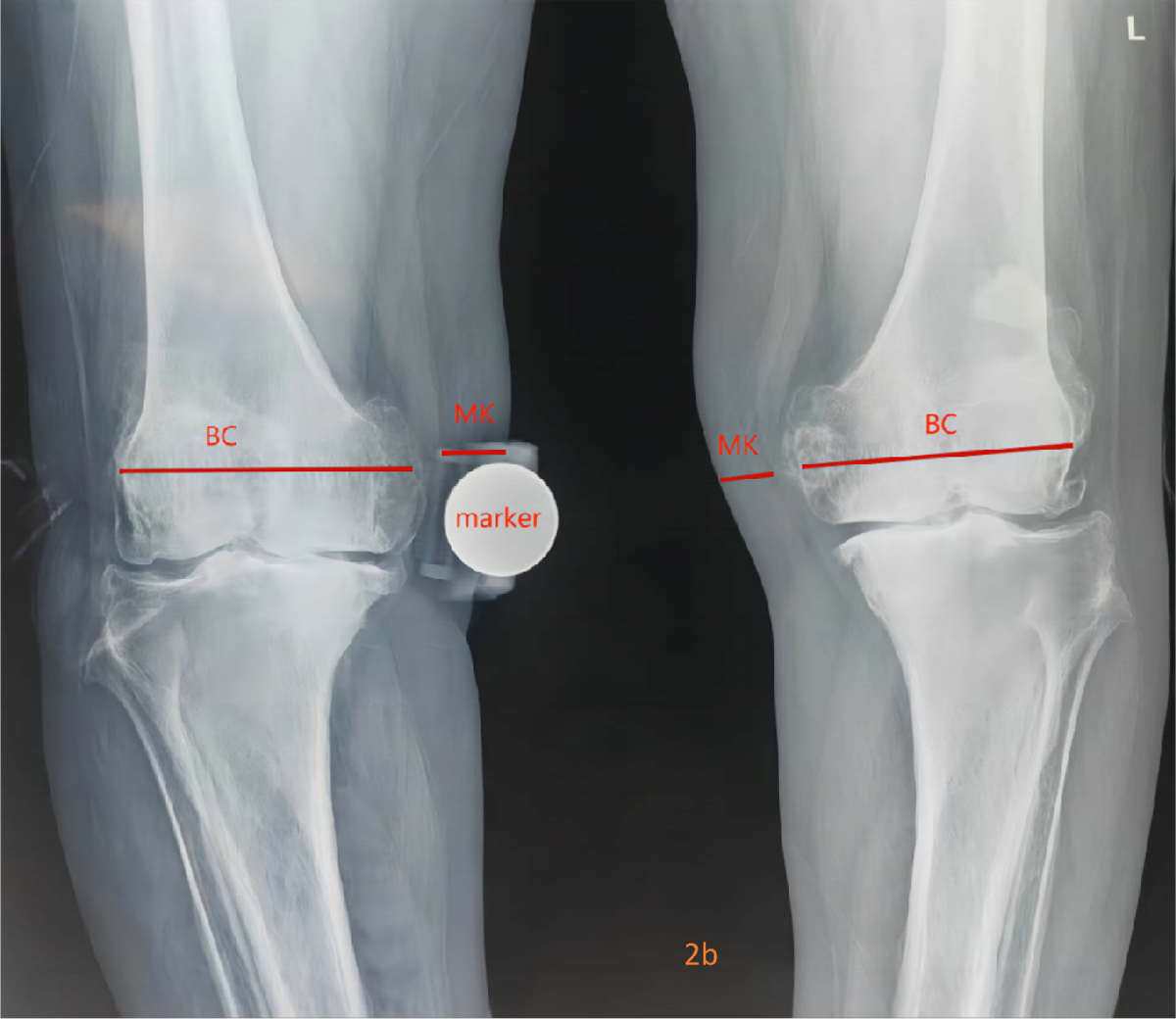
Measurement of soft tissue and bone thickness on X-ray film of knee joint Fig. 2a Measurement of soft tissue and bone thickness on lateral radiograph of knee joint AF Anterior femur Subcutaneous tissue: On the lateral film of the knee joint first locates the upper edge of the femoral condyle, 3cm upward along the upper edge of the femoral condyle, locates the measurement section, and measures the distance from the upper surface of the quadriceps femoris to the skin surface on the measurement section AT Anterior tibia Soft tissue: Distance from the surface of tibial tubercle to the skin AP Antepatellar Soft tissue: Distance from the midpoint of the anterior surface of the patella to the skin P Patella thickness: Solid distance between the midpoint of the anterior and posterior surfaces of the patella Fig. 2b Measurement of soft tissue and bone thickness on the anteroposterior radiograph of knee joint MK Medial knee Subcutaneous tissue: Measure the distance from the upper surface of the medial collateral ligament to the skin in the medial condyle of the knee joint BC Femoral condyle width: The line between the midpoint of the medial edge of the medial

### Evaluate

In order to determine whether the patient’s preoperative BMI and soft tissue indicators on preoperative anteroposterior and lateral views of the knee joint are associated with postoperative complications leading to readmission, we divided the 761 joint replacements into 22 cases with readmission and 739 cases without readmission complications. The IBM SPSS Statistics 26.0 system was used to perform independent sample T-tests on the two sets of data. First, the Levene test for homogeneity of variance was performed to determine whether the variances of the two sets of data were equal. If equal, a t-test was used. If the variances were different, a corrected t-test was used to compare whether there was a statistical difference between the two groups.

Finally, in order to determine the effect of soft tissue measurement indicators and whether they are associated with BMI and patient weight, patients were matched according to BMI and weight using PSM propensity score matching (1:n) for cases with readmission complications. The IBM SPSS Statistics 26.0 system was used to perform paired sample T-tests on the two sets of data to compare the differences between the sample group and the readmission complication group.

## Results

A total of 761 joint replacements from 625 patients were included in the study, of which 22 knee joints had readmission events due to postoperative complications. There were no differences in age, gender, presence of comorbidities, ASA score or side of surgery between the two groups.

In the study of readmission complications, there were statistically significant differences in all comparisons of periarticular soft tissue thickness defined by us between the complication group and the control group. However, regarding BMI between the two groups, the readmission complication group had a higher value of 28.36±3.38 compared to the non-readmission complication group of 27.15±3.63, but we did not find any statistical difference. No difference was found in weight between the two groups.

In the measurement of anteroposterior and lateral views of the joint, the antepatellar soft tissue of the readmission complication group was 8.72±5.63cm, compared to 7.05±2.97cm for the non-complication group (t=2.509, sig.(2-tailed)=0.012), showing a significant difference. In the comparison of anterior femur subcutaneous tissue thickness measurement, the readmission complication group was 13.55±5.67cm compared to 10.43±4.04cm for the non-complication group (t=3.455 sig.(2-tailed)=0.001). In the comparison of pretibial soft tissue thickness measurement, the readmission complication group was 10.30±7.26cm compared to 7.58±4.67cm for the non-complication group (t=2.664, sig.(2-tailed)=0.008). Both indicators showed significant differences. In the comparison of medial knee subcutaneous tissue thickness measurement, the readmission complication group was 23.47±8.30cm compared to 18.89± 6.71cm for the non-complication group (t =3.140 sig.(2-tailed)=0.002). In our set ratios, antepatellar soft tissue ratio is defined as antepatellar soft tissue thickness to patellar bone thickness ratio and medial knee subcutaneous tissue ratio is defined as medial knee subcutaneous tissue thickness to femoral condyle width ratio. The non-complication group had a ratio of 0.390±0.175 compared to 0.496±0.331 for the readmission complication group (t=2.706 sig.(2-tailed)=0.007). The non-complication group had a ratio of 0.228±0.0846 compared to 0.279 ±0.098 for the readmission complication group (t=2.799 sig.(2-tailed)=0.005). Both ratios showed significant differences between the two groups.

In the further matching test process, we used PSM propensity score matching to match the BMI and weight of patients with 22 knee joints with readmission events and 1:4 matched knee joints without readmission events. After matching, the two groups analyzed the indicators of periarticular soft tissue thickness. The results showed that in the comparison of anterior femur subcutaneous tissue thickness measurement, the readmission complication group was 13.55±5.67cm compared to 10.61 ± 4.15cm for the non-complication matched group (t=2.753 sig.(2-tailed)=0.007), still showing a significant difference. In the comparison of pretibial soft tissue thickness measurement, the readmission complication group was 10.30±7.26cm compared to 8.04±4.37cm for the non-complication matched group (t=1.886, sig.(2-tailed)=0.062). In the comparison of medial knee subcutaneous tissue thickness measurement, the readmission complication group was 23.47±8.30cm compared to 19.69±7.05cm for the non-complication matched group (t=1.970, sig.(2-tailed)=0.058). All of these soft tissue or subcutaneous tissue thicknesses showed general statistical differences between the two groups.

In our set ratios, antepatellar soft tissue ratio is defined as antepatellar soft tissue thickness to patellar bone thickness ratio and medial knee subcutaneous tissue ratio is defined as medial knee subcutaneous tissue thickness to femoral condyle width ratio. The non-complication group had a ratio of 0.410±0.167 compared to 0.496±0.331 for the readmission complication group (t=1.734 sig.(2-tailed)=0.086). The non-complication group had a ratio of 0.240±0.088 compared to 0.280 ±0.099 for the readmission complication group (t=1.706 sig.(2-tailed)=0.098). Both groups showed general statistical differences.

## Discussion

This study used clinical and imaging data from 761 joint replacements performed in 2019-2020 to show that measuring periarticular soft tissue indicators on anteroposterior and lateral views of the knee joint can better predict readmission complications and infection events in patients compared to BMI.

The study of whether obesity can be an independent risk factor for post-total knee arthroplasty has a long history. Jaiben George et al. (2014) found through a Nationwide Inpatient Sample study that the impact of BMI-defined morbid obesity on post-total knee arthroplasty seems to be quite mild and suggested that continued research is needed to determine the relationship between obesity and post-joint replacement complications [7]. Ali J. Electricwala, MD et al. found that early reinfection of TKA was significantly higher in those with increased BMI (54%, 74/138) than in those with normal BMI (24%, 8/33, P<0.003), but acute, early, mid or late revision TKA due to aseptic loosening and/or bone resorption, instability, stiffness or other reasons did not significantly increase [17]. In 2018, Kwasny MJ et al. constructed multiple models of BMI and found that the risk of post-joint replacement complications did not increase uniformly with increasing BMI and therefore further concluded that BMI cannot be used as an independent predictor of risk [18]. This result was also validated by Evans JT et al. in a large-scale study on survival rate in 2022. There was no evidence from large national registry statistics that high BMI patients had worse prognosis [19].

BMI is a good screening tool for obesity in the population, but it does not seem to be a good indicator for predicting the prognosis of total knee arthroplasty. It is worth noting that the factors that cause adverse effects after total knee arthroplasty due to obesity include local inflammation caused by fat accumulation, chronic inflammation and pain susceptibility caused by hyperlipidemia, biomechanical changes caused by high weight, and various complications caused by obesity [20–23]. Of these indicators, only higher weight may be closely associated with BMI, while the rest can be better evaluated individually independent of BMI.

Anteroposterior and lateral views of the knee joint are important imaging data for assessing the progression of osteoarthritis in patients [24]. However, orthopedic surgeons do not fully exploit the information on anteroposterior and lateral views of the knee joint and often focus on changes in bone information. The density differences between subcutaneous fat and muscle can also be reflected in increasingly refined X-ray imaging data [25]. Therefore, compared to past physical examinations, judging muscle strength and estimating BMI, doctors can now more specifically understand the patient’s subcutaneous fat and muscle conditions from X-rays. Jacob M. et al. studied four X-rays before total knee arthroplasty (TKA) in patients with body mass index (BMI)>30 kg/m2 and found that there were large differences in overall patterns of obesity among same-sex subjects with similar BMI and no direct connection between BMI and soft tissue on imaging [26]. Later, Russell A et al.’s study of 572 joint replacement patients showed that prepatellar fat thickness on lateral views of the joint can well assess infection at the surgical site [28], while Vikesh Kumar Gupta et al.’s study found that pretibial soft tissue is a protective factor for superficial infection and prepatellar soft tissue has no effect. However, 62 out of 494 patients in this experiment had superficial incision complications, which is much higher than the usual complication rate of joint replacement [28]. In 2020, Hamed Vahedi, MD et al.’s study found that differences in medial knee soft tissue thickness were associated with periprosthetic infection after surgery [29].

This retrospective study has many advantages. First, in terms of patient sample selection, patients collected from 2019-2020 and aged 65-80 years underwent standard knee replacement surgery and perioperative treatment. The surgeons were all senior orthopedic doctors with more than 200 joint replacement surgeries, minimizing bias caused by different treatments. Second, in terms of data collection, BMI and various soft tissue data on anteroposterior and lateral views of the joint mentioned in previous studies were integrated. Finally, in terms of experimental analysis, independent sample tests were first used to determine indicators related to post-joint replacement prognosis, and then matching was used to eliminate the influence of BMI and other factors, providing ideas for further exploration of postoperative management of patients. However, this study also has many limitations. As a retrospective study, it has its inherent shortcomings. Second, in order to ensure that patients receive similar and mature surgical treatment and for the convenience of data collection, the data used in this experiment are from multiple centers in a single hospital and the subjects are mainly Asians. The number of morbidly obese (BMI>40) patients may be relatively small. The patient data collected is X-ray imaging, which is relatively rough compared to CT and MRI for judging soft tissue thickness. Finally, the knee joint flexion angle controlled in the anteroposterior and lateral views of the knee joint collected in this experiment is 10°-30°. However, many medical institutions may not meet this standard when taking anteroposterior and lateral views of the knee joint. Overly flexed knee joints may contract prepatellar soft tissue and pretibial soft tissue. Therefore, there are requirements for the quality of images in practical applications.

## Conclusion

In summary, as a risk factor for total knee arthroplasty, obesity has not shown good efficacy in evaluating the association between patient obesity and adverse events after TKA using BMI. Preoperative measurement of soft tissue data on anteroposterior and lateral views of the patient’ s knee joint can better predict readmission complications after total knee arthroplasty. Thicker periarticular soft tissue may be associated with a higher risk of readmission after surgery.

## Data Availability

All data produced in the present study are available upon reasonable request to the authors

---

**Table 1.** Baseline Characteristics of the Study Population.

| Baseline Characteristics of the Study Population |  |  |  |
| --- | --- | --- | --- |
|  | Events of readmission Complication |  | P Value |
|  | Yes (N=22) | No (N=739) |  |
| Age |  |  |  |
| 65-70 | 10 | 290 | 0.593 |
| 71-75 | 7 | 315 |  |
| 76-80 | 5 | 134 |  |
| Gender |  |  |  |
| female | 19 | 579 | 0.367 |
| male | 3 | 160 |  |
| Diabetes |  |  |  |
| yes | 18 | 657 | 0.301 |
| no | 4 | 82 |  |
| ASA physical status |  |  |  |
| Class 1 | 4 | 90 | 0.665 |
| Class 2 | 18 | 645 |  |
| Class 3 | 0 | 4 |  |
| Class 4 | 0 | 0 |  |
| Surgical side |  |  |  |
| left | 10 | 369 | 0.67 |
| right | 12 | 370 |  |

**Table 2.** Multiple independent sample t-test of readmission complications.

| Multiple independent sample t-test of readmission complications |  |  |  |
| --- | --- | --- | --- |
|  | Events of readmission Complication |  | P Value |
|  | Yes (N=22) | No (N=739) |  |
| **Weight , kg** | 72.35±11.70 | 69.67±11.16 | 0.301 |
| **BMI , kg/m2** | 28.36±3.38 | 27.17±3.63 | 0.111 |
| **Soft tissue thickness , mm** |  |  |  |
| Antepatellar Soft tissue | 8.72±5.63 | 7.05±2.97 | 0.012 |
| Anterior femur Subcutaneous tissue | 13.55±5.67 | 10.46±4.09 | 0.001 |
| Anterior tibia Soft tissue | 10.30±7.26 | 7.56±4.67 | 0.008 |
| Medial knee Subcutaneous tissue | 23.47±8.30 | 18.89±6.70 | 0.002 |
| **Soft tissue to bone ratio** |  |  |  |
| Antepatellar Soft tissue to Patellar ratio | 0.496±0.331 | 0.390±0.175 | 0.007 |
| Medial knee tissue to Femoral condyle ratio | 0.280±0.099 | 27.15±0.085 | 0.005 |

**Table 3.**
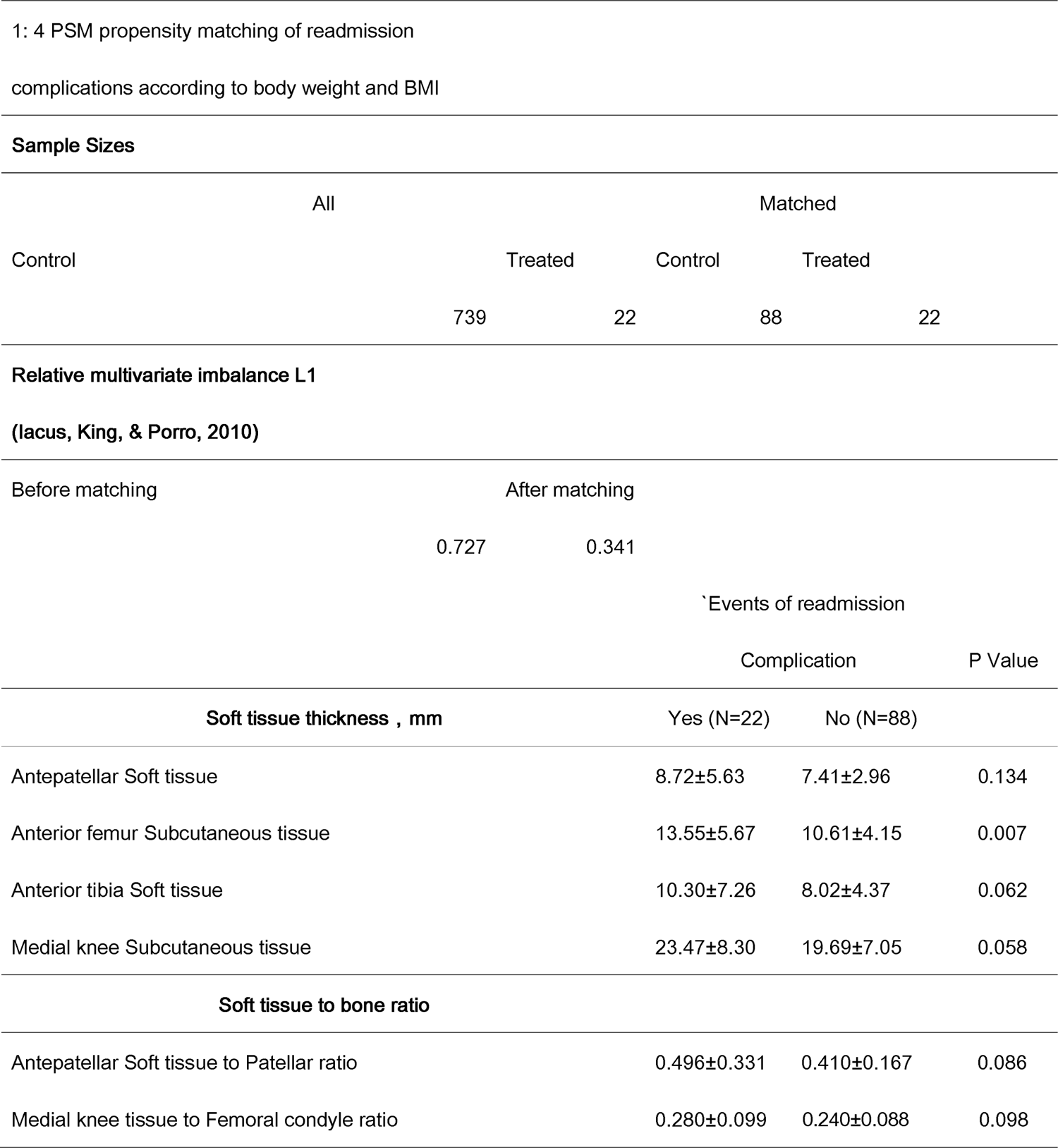
1: 4 PSM propensity matching of readmission complications according to body weight and BMI.

